# The relationship between new PCR positive cases and going out in public during the COVID-19 epidemic in Japan

**DOI:** 10.1101/2021.03.07.21252959

**Authors:** Hiromichi Takahashi, Iori Terada, Takuya Higuchi, Daisuke Takada, Jung-ho Shin, Susumu Kunisawa, Yuichi Imanaka

## Abstract

Suppression of the first wave of COVID-19 in Japan is assumedly attributable to people’s increased risk perception by acquiring information from the government and media reports. In this study, going out in public amidst the spread of COVID-19 infections was investigated by examining new polymerase chain reaction (PCR) positive cases of COVID-19 and its relationship to four indicators of people going out in public (the people flow, the index of web searches for going outside, the number of times people browse restaurants, and the number of hotel guests), from the Regional Economic and Social Analysis System (V-RESAS). Two waves of COVID-19 infections were examined with cross-correlation analysis. In the first wave, all four indicators of going out reacted oppositely with the change in new PCR positive cases, showing a lag period of –1 to +6 weeks. In the second wave, the same relationship was only observed for the index of web searches for going outside. These results suggest that going out in public could not be described by new PCR positive cases alone in the second wave, even though they could explain people going out to some extent in the first wave.

## 1. Introduction

Novel coronavirus disease 2019 (COVID-19), caused by infection with severe acute respiratory syndrome coronavirus two (SARS-CoV-2), which can lead to severe pneumonia in infected humans, emerged as an unprecedented global pandemic in early 2020. After the first outbreak in China in December 2019, the disease has continued to spread throughout the world and has had a considerable impact on everyday life [1-2]. In Japan, the first case of COVID-19 infection was confirmed on January 16 [3], and since then, the number of cases and deaths has continuously fluctuated. The first wave of COVID-19 infections was defined as occurring from March to May 2020 [4].

The government administered various policies to influence people flow control and expansion in response to the spread of infection. For instance, the government designated COVID-19 equivalent to a category two infectious disease [5-6] and postponed the Olympic games [7]. In addition, the government requested various events be canceled [8] and that schools and high-risk facilities, such as pubs, be closed [9-10]. A state of emergency was declared [11-15], and people were asked to refrain from going out in public to reduce person-to-person transmission by 70%–80% [16]. These measures were requested of the public but were not compulsory, and there were no penalties for disregarding these initiatives. Nevertheless, people followed the requests and refrained from going out in public to a certain extent.

When the first wave was restrained, the government proposed a new public lifestyle to prevent the spread of infection. The guidance stipulated avoiding the “three Cs”: closed spaces, crowded places, and close contact settings [17]. Furthermore, the government and each prefecture monitored several infection indicators and set specific standards to combat subsequent infections [18-20]. These measures were intended to minimize the risk of repeating the negative impacts of the first wave from a long-term perspective while keeping in mind the need to coexist with the virus. As an economic measure, the government provided 100,000 yen per person during the first wave [21]. However, the damage to the economy was tremendous [22] due to people’s self-restraint in avoiding going out to suppress the first wave of the pandemic. Several stimulus measures were implemented after the first wave, including subsidies for travel and restaurant industries [23-28]. The economic stimulus measures were designed to encourage people to go out and move around while the infection was spreading, and it could be inferred that this increased the number of people traveling and eating out.

As previous studies have shown, the end of the first wave was achieved by people’s self-restraint from going out in public, which was attributed to increased public awareness and understanding of the risks of the current situation through media reports [29-34]. In addition, the characteristics of Japanese culture may have contributed to the lower number of new polymerase chain reaction (PCR) positive cases and deaths per population compared to other countries [35]. Watanabe et al. attributed the decrease in the number of people going out in public during the first wave to government announcements, such as daily news releases of new PCR positive cases [36]. Meanwhile, anxiety and precautionary behaviors decreased as people become more aware of their subjective risk perception as an immediate threat [37]. However, differences in going out behaviors between the first and subsequent waves in Japan have not been clarified. Daily news reports, including new PCR positive cases, informed the public of the seriousness of the COVID-19 epidemic. Based on the available information of the situation, and including both objective and subjective reasoning, people then decide how they will behave, for example, by choosing whether to go out in public. The information reported in the media is expected to influence individual behavior and either discourage or encourage going out (especially concerning the three Cs that are considered high risk).

In Japan, the government and prefectures have set up indicators of public movements that should be monitored [18-20]. Nakano et al. also developed a new “K-value” indicator which they insist could predict the prevalence of infection [38]. The K-value and many other indicators set by the prefectures rely on daily reports of new PCR positive cases. Furthermore, according to Watanabe et al., new infections resulted in people refraining from going out in public [36]. However, the difference in people going out between the first and subsequent waves has not been clarified. Thus, this study investigated the lag between going out behaviors in response to the spread of infection by examining the relationship between the new PCR positive cases and each of four indicators of going out: 1) the people flow at specific locations in Japan; 2) the index of web searches for going outside; 3) the number of times people browse restaurants; 4) the number of hotel guests. By examining changes in people going out in public between the first and subsequent waves, and the underlying changes in people’s awareness of the crisis as well as the effects of government policies, this study will provide insight for implementing effective countermeasures against the spread of infection.

## 2. Materials and Methods

### 2-1. Data Collection

#### 2-1-1. New PCR positive cases

New PCR positive cases of COVID-19 are announced daily by the Ministry of Health, Labor, and Welfare. This number is based on the date each prefecture receives a positive case report from medical institutions, not necessarily the date of infection onset. The data was obtained from “Toyo Keizai Online”, a Japanese publication that independently compiles data from the Ministry of Health, Labor, and Welfare [39]. The published data is an indicator for implementing various policies and informing the news that people see every day, thereby influencing public awareness. The number of cases is tallied weekly, Monday through Sunday.

#### 2-1-2. Four indicators of going out in public

The Regional Economic and Social Analysis System (V-RESAS) website reports four public movement indicators based on data provided by the Office for Promotion of Regional Revitalization, Cabinet Office, Government of Japan [40]. Data from multiple sources is compiled, and V-RESAS shows the cumulative outcomes, showing weekly change rates compared to the same period last year [40].

##### 2-1-2-1. The people flow

The people flow shows the weekly change in the rate of people moving through several locations in Japan compared to the same week of the previous year. Agoop is a company that collects location information derived from their own applications and other companies that allow Agoop to use their location data [40,41]. Agoop’s data is used by the V-RESAS and various media outlets, including the Japanese public broadcaster NHK [42].

##### 2-1-2-2. The index of web searches for going outside

The index of web searches for going outside was derived from Yahoo Japan Corporation’s data, which uses AI technology to categorize words entered into Yahoo Search [40,43].

##### 2-1-2-3. The number of times people browse restaurants

The number of times people browse restaurants was derived from restaurant information views on the Food Data Platform provided by Retty. It is a large-scale platform of food businesses that has 40 million monthly users [40,44].

##### 2-1-2-4. The number of hotel guests

Accommodation data from the Tourism Forecast Platform, the secretariat of the Japan Travel and Tourism Association, informed the number of guests staying at hotels. The anonymized data was collected from travel agency storefronts and reservation sites, and included more than 130 million stays (as of September 2020) [40,45].

### 2-2. Definitions of COVID-19 epidemic periods

In this study, COVID-19 infections in Japan were divided into two periods: the first wave (from January 16, 2020, when the first infected person was identified, to the end of May [4]), and the second wave (from June 1, 2020, to the first week of November).

### 2-3. Analysis

Cross-correlation analysis was performed to investigate the relationship between new PCR positive cases and each of the four indicators of going out in public for each of the first and second waves. The cross-correlation function (CCF) describes the relationship between two time-series datasets, X(t) and Y(t), and has been used as a method to estimate the time lag between the datasets. One infectious disease study previously used this method for an outbreak of acute exanthematous illness attributed to Zika virus, Guillain-Barre syndrome, and microcephaly in Brazil during 2014–2015 [46]. In addition, CCF was used to estimate the speed of influenza epidemics by comparing lag in drug sales between pharmacies that were geographically separated in Japan [47]. Moreover, it has been used to detect the correlation pattern between output and nominal variables in economics [48]. Following the previous study [47], CCF was defined as:^1)^

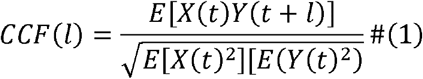

where *CCF*(*l*) denotes the cross-correlation function between two time series, *X*(*t*) and *Y*(*t*), E[.] denotes the mean over time *l* of the random variables inside the square brackets, and *l* denotes the lag.

Interpreting the results of CCF analysis focuses on two points: the sign of the lag and the sign of the cross-correlation. For the former, if the lag is negative, the time series data X(t) is ahead of Y(t). On the other hand, if the lag is positive, the time series data X(t) can be expressed as lagging behind Y(t), i.e., the lag is judged by whether it is to the right side (positive region) or the left side (negative region). For the latter, when the cross-correlation is positive, the forms of the two indicators are similar, and when it is negative, the forms of the two indicators are different. In other words, when cross-correlation is positive, the result shows how much lag there is between the two indicators, and when it is negative, the result shows how much lag there is between the change in X(t) and the opposing change in Y(t). Therefore, the sign of the lag and that of the cross-correlation for each combination of indicators was evaluated in this study. Moreover, significance in the negative region indicates that new PCR positive cases precede the other indicators, while significance in the positive region indicates that the other indicators precede new PCR positive cases.

Statistical significance was regarded as a two-sided P-value <0.05. All analyses were performed using R 3.6.3 (R Foundation for Statistical Computing, Vienna, Austria) and the astsa package (version 1.10).

Any approvals were not needed, since we used only open data in this study, which can not identify any individuals.

## 3. Results

The timeline of changes in new PCR positive cases, the indicators of going out in public, and representative policies are shown in Figure 1.

**Figure 1.**
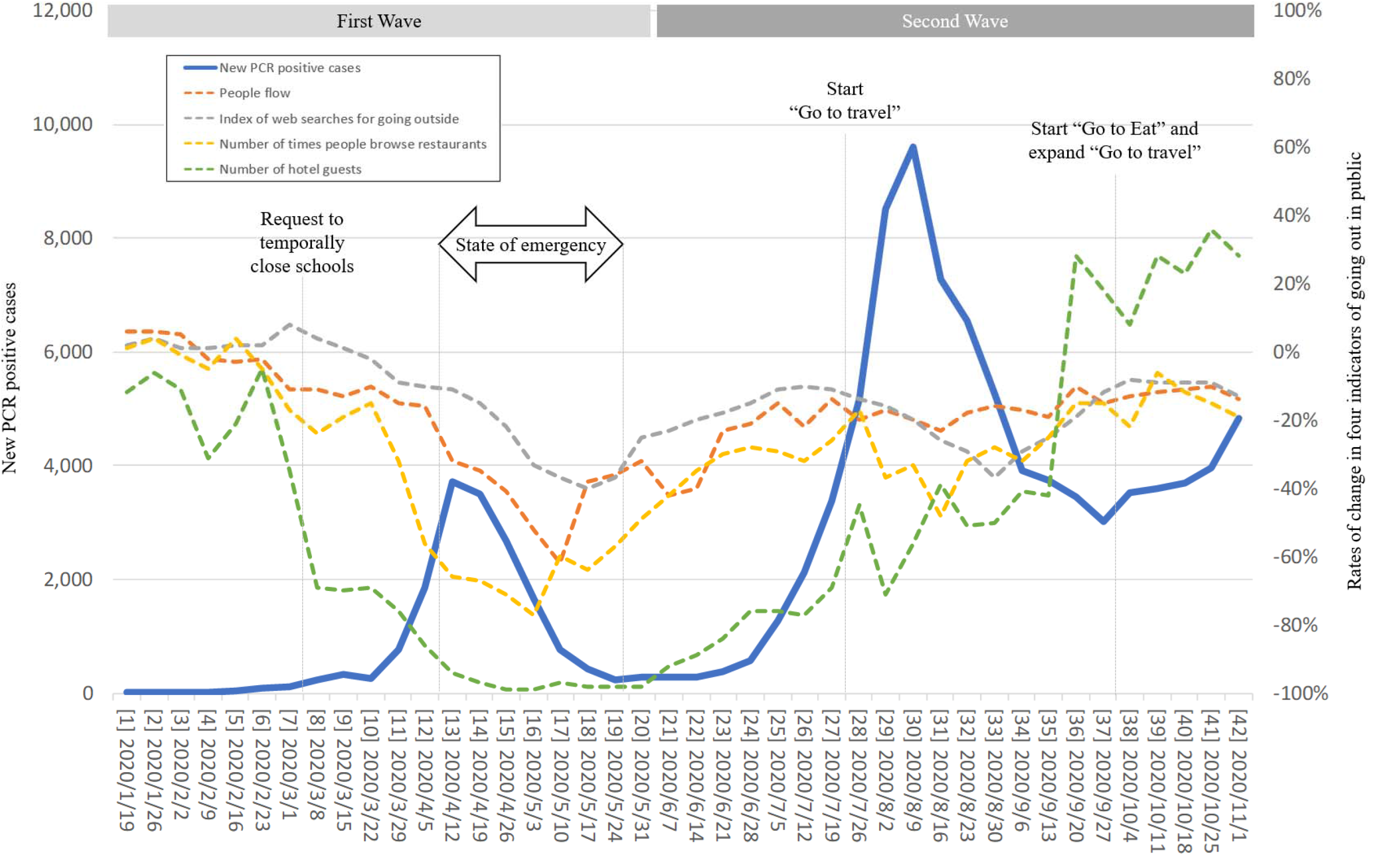
Time trend of new PCR positive cases, four indicators related to going out in public, and representative policies. Each number before the date on the x-axis is the consecutive number of weeks, with the week of January 19, 2020, the starting date of the data series, being 1.

For the first wave, the cross-correlation coefficients of new PCR positive cases and people flow were negative in the lag region –4 to 0 (Figure 2A). For new PCR positive cases and the index of web searches for going outside, cross-correlation coefficients were significantly negative in the lag region –6 to –1 (Figure 2B). For the number of times people browse restaurants, cross-correlation coefficients with new PCR positive cases were negative between lag –4 to +1 (Figure 2C). Finally, cross-correlation coefficients for the number of hotel guests and new PCR positive cases were negative between lag –3 to +1 (Figure 2D) and were also positive in the lag region +10 to +12 (Figure 2D). Therefore, the results show that new PCR positive cases preceded people flow and the index of web searches for going outside. Similarly, new PCR positive cases preceded (or coincided with) the number of times people browse restaurants and the number of hotel guests. Moreover, new PCR positive cases and the other four indicators had opposite forms since the cross-correlation coefficients were negative for all combinations of indicators. Incidentally, for the last combination of indicators, the reasons why the observed lags in the positive region can be ignored were described in the discussion section.

**Figure 2.**
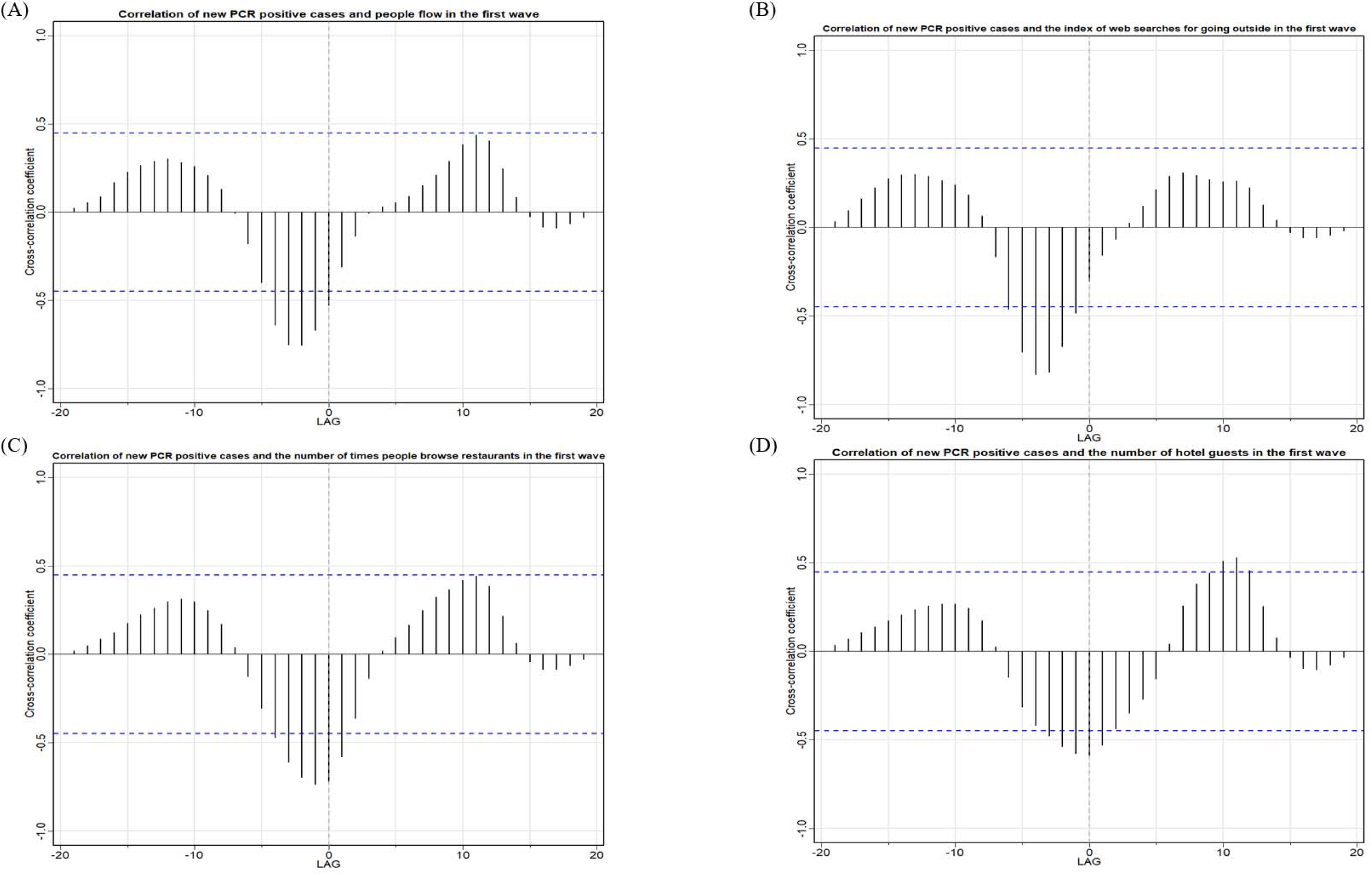
Cross-correlation of new PCR positive cases with A) people flow, B) the index of web searches for going outside, C) the number of times people browse restaurants, and D) the number of hotel guests in the first wave. Dashed blue lines indicate 95% confidence intervals for a null model of no association.

The second wave results were complicated compared to the first wave because the cross-correlation coefficients were negative, neutral (0), or positive, and the lag times were significant in the negative or positive regions, depending on the combination of indicators (Figure 3).

**Figure 3.**
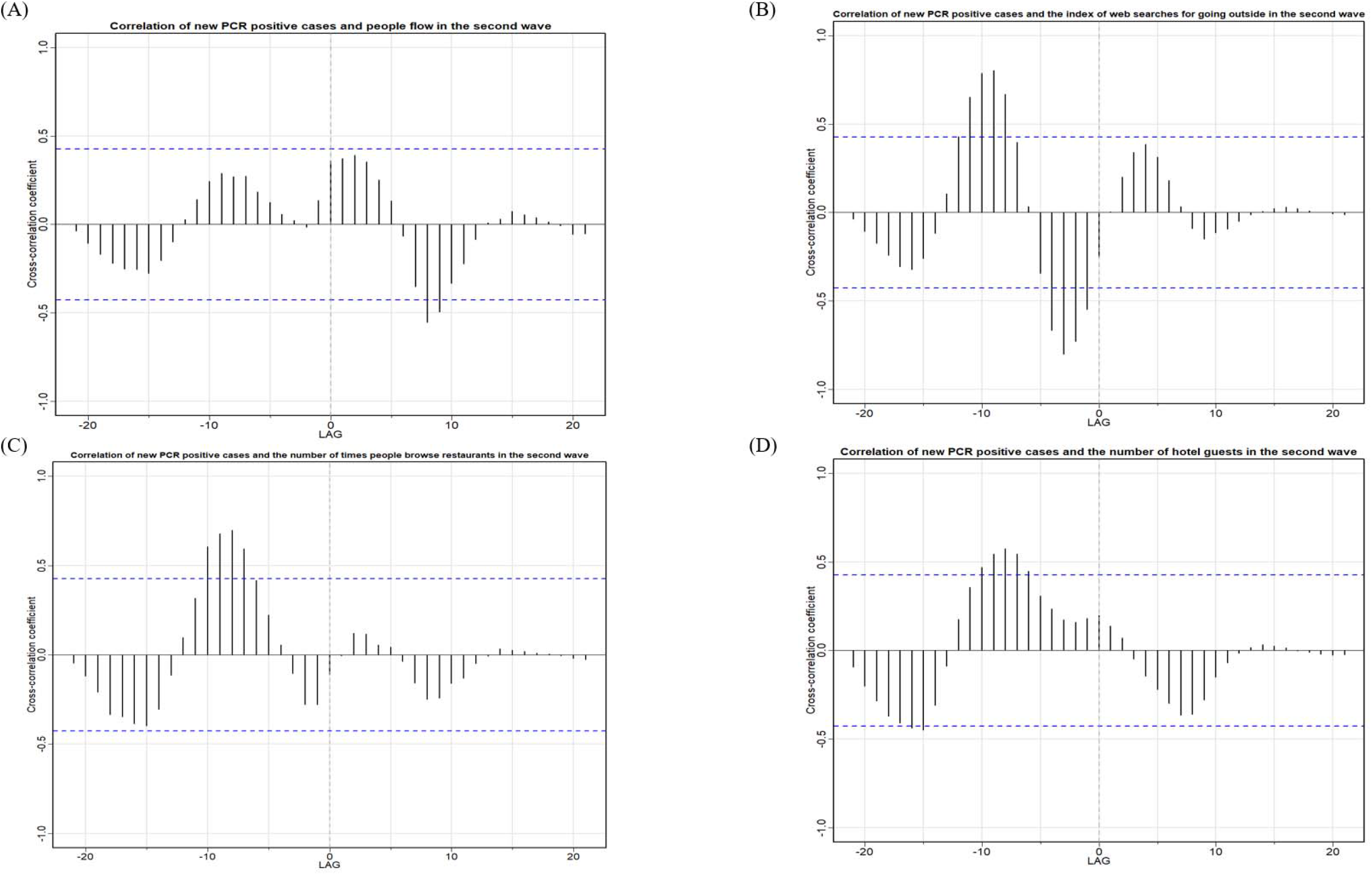
Cross-correlation between new PCR positive cases with A) people flow, B) the index of web searches for going outside, C) the number of times people browse restaurants, and D) the number of hotel guests in the second wave. Dashed blue lines indicate 95% confidence intervals for a null model of no association.

The cross-correlation coefficients of new PCR positive cases and people flow were significantly negative in the lag region +8 to +9 (Figure 3A), which means that the peak in new PCR positive cases lagged behind people flow (i.e., the peak in new PCR positive cases came after the people flow). This result was partially opposite to the first wave, in which new PCR positive cases preceded people flow.

Regarding the index of web searches for going outside, significant positive and negative cross-correlation coefficients with new PCR positive cases were found in distinct areas of the negative lag region (Figure 3B). Firstly, cross-correlation coefficients were significantly negative in the lag region –4 to –1 (Figure 3B). This outcome confirms a similar structure for this combination of indicators to the first wave. In other words, the peak of new PCR positive cases preceded the nadir of the index of web searches for going outside, and these two indicators were different. Secondly, cross-correlation coefficients were significantly positive in the lag region –12 to –8 (Figure 3B). The peak (or nadir) in new PCR positive cases preceded the peak (or nadir) of the index of web searches for going outside, and the forms of the two indicators were similar. Such a structure was not observed in the first wave.

For the number of times people browse restaurants, cross-correlation coefficients with new PCR positive cases were significantly positive in the lag region –10 to –7 (Figure 3C). Therefore, it showed that the peak (or nadir) of new PCR positive cases preceded the peak (or nadir) of the number of times people browse restaurants and that the two indicators were similar. This result was partially opposite to the first wave because the two indicators differed in the first wave.

The number of hotel guests had significant positive and negative cross-correlation coefficients with new PCR positive cases in distinct negative lag regions. Firstly, the cross-correlation coefficients were significantly positive in the lag region –10 to –6 (Figure 3D). The peak (or nadir) of new PCR positive cases preceded the peak (or nadir) of the number of hotel guests, and the forms of the two indicators were similar. This result was partially opposite to the result of the first wave because the forms of the two indicators were different in the first wave for these indicators. Secondly, the cross-correlation coefficients were significantly negative in the lag region –16 to –15 (Figure 3D). The peak in new PCR positive cases preceded the nadir of the number of hotel guests, and these two indicators were different.

## 4. Discussion

In this study, we analyzed the change in new PCR positive cases with four indicators about people going out in public during the first and second waves of COVID-19 infection in Japan. People flow, the index of web searches for going outside, the number of times people browse restaurants, and the number of hotel guests reacted with similar timing to the change in new PCR positive cases in the first wave. On the other hand, the same reaction was only observed for the index of web searches for going outside in the second wave.

### 4-1. The first wave

All four indicators responded significantly in the negative lag region at or near the same time as the change in new PCR positive cases. It has been reported that lockdown effectively controls the spread of infection [49], but in Japan, the government only asked people to abstain from going out in public voluntarily. Nevertheless, people followed the government’s request to refrain from going out to protect themselves from the unknown virus by limiting their contact with others. During the first wave, new information about the coronavirus continued to be disseminated daily, along with reports of new PCR positive cases. It is assumed that this information gave people a sense of urgency and encouraged them to refrain from going out. This is consistent with previous research that showed risk perception increased in the early stages of the epidemic, and self-restraint became widely practiced [29-36].

The indicators of going out in public had various lag times from new PCR positive cases. The starting point and the peak in significant lag time for the number of times people browse restaurants and the number of hotel guests were relatively earlier than the people flow and the index of web searches for going outside. This observation suggests that people were particularly aware of the risk of eating out and staying at hotels, and they refrained from these activities accordingly. Furthermore, this outcome is assumed to result from the request by government and local authorities for people to refrain from traveling and eating out and because public transport, hotels, and restaurants were considered dangerous at the beginning of the outbreak.

As for the positive lag between new PCR positive cases and the number of hotel guests (lag +10 to +12; Figure 2D), this result is likely due to the data in the period before the COVID-19 epidemic occurred.

### 4-2. The second wave

There were two characteristic reactions in the second wave. Comparing to the first wave, a different reaction was observed for people flow, the number of hotel guests, and the number of times people browse restaurants. In addition, a significant positive response was seen in the number of hotel guests, the index of web searches for going outside, and the number of times people browse restaurants six to twelve weeks after the new PCR positive cases fluctuated.

For the first characteristic, there are three possible reasons why the second wave’s results differed from the first wave, except for the index of web searches for going outside. Firstly, a state of emergency was not declared in the second wave. In addition, the messages from the government in the second wave were not as strong and directive as compared to the first wave. Secondly, there was a positive effect of the “Go To Travel and Eat” campaign that encouraged people to go out. Although not statistically significant, the number of hotel guests showed a positive trend immediately after the change in new PCR positives cases, suggesting that the “Go To Travel” campaign effectively promoted travel. There was little negative change in the number of times people browse restaurants immediately after the change in new PCR positive cases, which may be because the “Go To Eat” campaign started in October while the “Go To Travel” campaign started on July 22. The timing of the change in the number of times people browse restaurants was late compared with the number of hotel guests [25-28]. Thirdly, people’s experiences in the first wave, the “new lifestyle” advocated by the government at the end of the first wave and the two factors mentioned above provided people with a vanity that spreading COVID-19 will calm down soon even when the infection spreads again. This result was consistent with the observation that people’s anxiety and preventive behaviors decrease as their risk perception of the seriousness as an immediate threat was reduced, as shown by Jones et al. [37]. The factors that encouraged people to stay at home during the first wave [29-36] have been recognized to some extent, and suggest that the sense of a crisis was not perceived beyond the first wave. In the second wave, the only change similar to that in the first wave was the index of web searches for going outside. However, the response to the change in new PCR positive cases was faster and ended earlier than in the first wave. This finding may have resulted from people knowing how to coexist with the virus better than during the first wave. Besides, while the index of web searches for going outside was decreasing, the people flow was not significantly reduced, suggesting that people were gradually resuming their public movements that did not require web searches, such as commuting and daily routine travel.

For the second characteristic in the second wave, three reasons can be attributed to the observations: 1) The “Go To Travel and Eat” campaign was expanded six to twelve weeks after the rapid increase in new PCR positive cases; 2) Almost all positive cases had recovered, and new PCR positive cases were nearing their nadir at the same time; 3) People might have become complacent about the pandemic.

### 4-3. Limitations

The data were sourced from the V-RESAS website. Since the data was from private entities, it does not include data from all people. However, the data used in this study is also used by policymakers and the media, so it is considered to have some validity [42]. There might be seasonal variations in people’s behavioral changes. Since the analysis period was from January 16 to the first week of November, the results might have been underestimated or overestimated because of seasonal variations that might not have been considered in the analysis. However, since people’s behavior during a pandemic differed significantly from a usual year, this limitation is not considered a serious problem.

## 5. Conclusions

In conclusion, people paid close attention to the ever-changing number of new PCR positive cases and refrained from going out in public during the first wave. However, in the second wave, people did not refrain from going out even though new PCR positive cases were more extensive than in the first wave. This finding suggests that going out in public was not explained by daily reports of new PCR positive cases alone. In the case of a possible future spread of the disease, it will be necessary to consider other factors that influence public awareness.

## Data Availability

The datasets generated during and/or analyzed during the present study are available from the
corresponding author on reasonable request.

## Acknowledgements

None

